# HRSA Population Health Dashboard

**DOI:** 10.1101/2023.04.17.23288701

**Authors:** Chi Nguyen

## Abstract

**Project Purpose:** To develop a quality dashboard tool for the Health Resources and Services Administration (HRSA) community health centers to examine health center characteristics associated with performance excellence and predict future healthcare cost.

**Background:** HRSA is the primary federal agency for improving health care to people who are geographically isolated, economically or medically vulnerable. HRSA-funded health centers are required to upload to Uniform Data System (UDS). HRSA health data is rich; however, their website provides limited meaningful visualization and data-driven analysis.

**Method:** The project reviewed data from Uniform Data System between 2017 and 2019. The data was cleaned using Excel. Visualization was displayed on Tableau, and the cost prediction modeling was completed using Python Machine Learning libraries. All the findings are available for public view on Weebly-hosted website.

**Outcomes:** The Tableau dashboard showcases the effectiveness of the preventive intervention and chronic disease management initiatives over the years. This tool focuses on comparison between outcomes in health care programs and cost benefits in all states. The dashboard also uses the most fit Machine Learning model and the most important variables to predict the cost per patient of a health center.

**Discussion:** The lack of similar performance comparison dashboards by HRSA suggests a need for HRSA Population Health Dashboard to understand the performances of HRSA-funded health centers or regions through times. It shows the ineffectiveness of the current healthcare programs in reducing disease growth and healthcare cost over the course of 3 years.

**Next Steps/Opportunities:** The final dashboard will be available for public use and will continue to aggregate more real-time data using Application Programming Interface (API) to improve its accuracy of prediction model. The HRSA Population Health Dashboard will be beneficial to management at both HRSA federal- and health center-level to make valuable strategies.

## Background

Health Resources and Services Administration (HRSA) is the primary U.S. federal agency for improving health care through its 90-plus programs and more than 3000 grantees to serve tens of millions of Americans who are geographically isolated, economically, or medically vulnerable (HRSA, 2020). Over 1400 health centers participate in HRSA’s clinical quality programs with the goal of delivering high-quality, affordable, and cost-effective health care.

Each year, HRSA-funded health centers and other entities receiving federal funding authorized under Section 330 of the Public Health Service Act are required to report their annual data to Uniform Data System (UDS) (HRSA, 2021). Included in the UDS are data on patient demographics, performance in clinical quality measures, staffing and service utilization, and financial data, as well as the number of physicians, certified nurse practitioners, and physician assistants, on-site or with whom the health center has contracts (Topmiller et al., 2020). UDS website provides detailed manuals and resource guidelines for health centers to standardize their data reporting (HRSA, 2021).

### Statement of Problem

With a concrete reporting system by UDS and extensive nationwide network of operation, HRSA has a rich source of health data. However, their downloadable files contain limited data. The rest of the data is only viewable on their website by going through the report of each health center. This restriction led to a limited number of visualization and data-driven analysis on HRSA healthcare performances.

### Literature Review

HRSA community health centers and UDS data have been used in several published studies and visualization galleries for performance analysis purpose.

HRSA has a set of dashboards available on their website focusing on COVID-19 operations, healthcare workforce and training program, demographics, Federal Tort Claims Act coverage, National Health Service Corps participation, organ donation and transplantation, medically underserved capacity, and health center operations (HRSA, n.d.). There is only one performance-focused dashboard on hypertension control at health center- and state-level.

Tableau Public is the most popular open platform to explore, create, and publicly share data visualizations among professionals. There are 34 dashboards on HRSA topic, and none of them emphasized quality performances (Tableau, n.d.).

Among traditional literature on HRSA quality improvement, Hu et al. (2018) evaluated the degree of improvement in health care quality in health centers with and without Patient-centered Medical Home (PCMH) recognition by using data from the 2012 to 2015 Uniform Data System and the Area Resource File. The research used descriptive analysis to look at the number differences in characteristics and clinical performances between PCMH grantees and non-PCMH grantees over the length of their PCMH implementation (Hu et al., 2018).

On the other hand, Lê-Scherban et al. (2019) examined the associations of hypertension and diabetes with nine measures of neighborhood socioeconomic status (poverty, education, deprivation index), social environment (violent crime, perceived safety and social capital, racial segregation), and built environment (land-use mix, intersection density). They used UDS data for quality measures and other data sets such as EHR data, American Community Survey, Philadelphia Police Department, and Southeastern Pennsylvania Household Health Survey to support their statistical analysis.

The literature review indicated available but limited resources and studies on HRSA comprehensive quality analysis at state-, city-, and health center-level.

### Project Purpose

The purpose of this project was to provide a data-driven tool for the HRSA community health centers to examine their characteristics associated with performance excellence over the years. It also consolidated full HRSA data for trendline and pattern analysis. Finally, it used Machine Learning to predict the healthcare cost of a health center provided other measures.

## Method

The project used HRSA health center data from years 2017 to 2019. Since downloadable HRSA public files did not include data for each health center, a Python script was used to scrape the data of health centers from HRSA website page by page. The final data set contained measures of age/ethnicity, characteristics, clinical, cost, and national program data. Measures with more than 500 empty records from health centers were removed. Data of 9 territories was excluded due to the low number of reported measures. Any health centers with missing data were eliminated from the measure aggregations and reporting. As shown in Figure 1, the remaining data set for visualization contained 29 measures and 1144 health centers for a period of 3 years.

**Figure 1.**
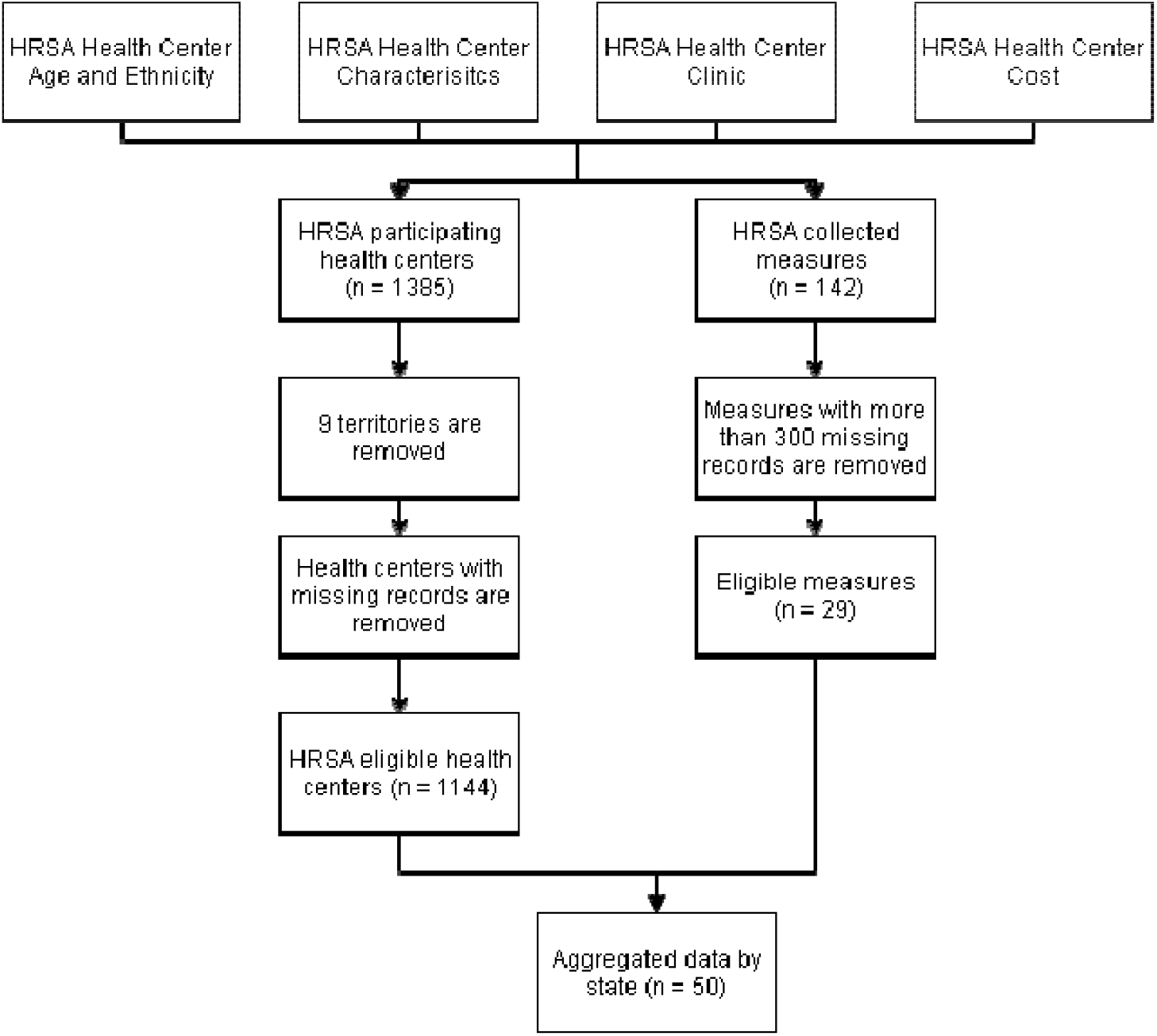
HRSA Data Cleaning Process

The cleaned data was then applied to produce data visualization and cost prediction model (see Figure 2). The data visualization was created using Tableau Desktop version 2020.2.12 and published to Tableau Public. The cost prediction was modelled with Python machine learning libraries. The data set was split into 70% for training and 30% for testing using 10-folds cross validation method. Each fold had different samples that were not present in other folds. This approach allowed thorough trainings on different samples in the dataset (Ilango, 2018). The following regression algorithms were tested to find the one with the best performance: Linear Regression, Lasso, Elastic Net, K Neighbors, Decision Tree, Support Vector Regression (SVR), Ada Boost, Random Forest, Extra Trees, and Gradient Boosting. The most important features for prediction were assessed and selected for the model. After the model was trained, it was applied to create an interactive cost prediction application using Streamlit application framework and Heroku cloud platform. Both the visualization and cost prediction app were hosted on a website powered by Weebly.

**Figure 2.**
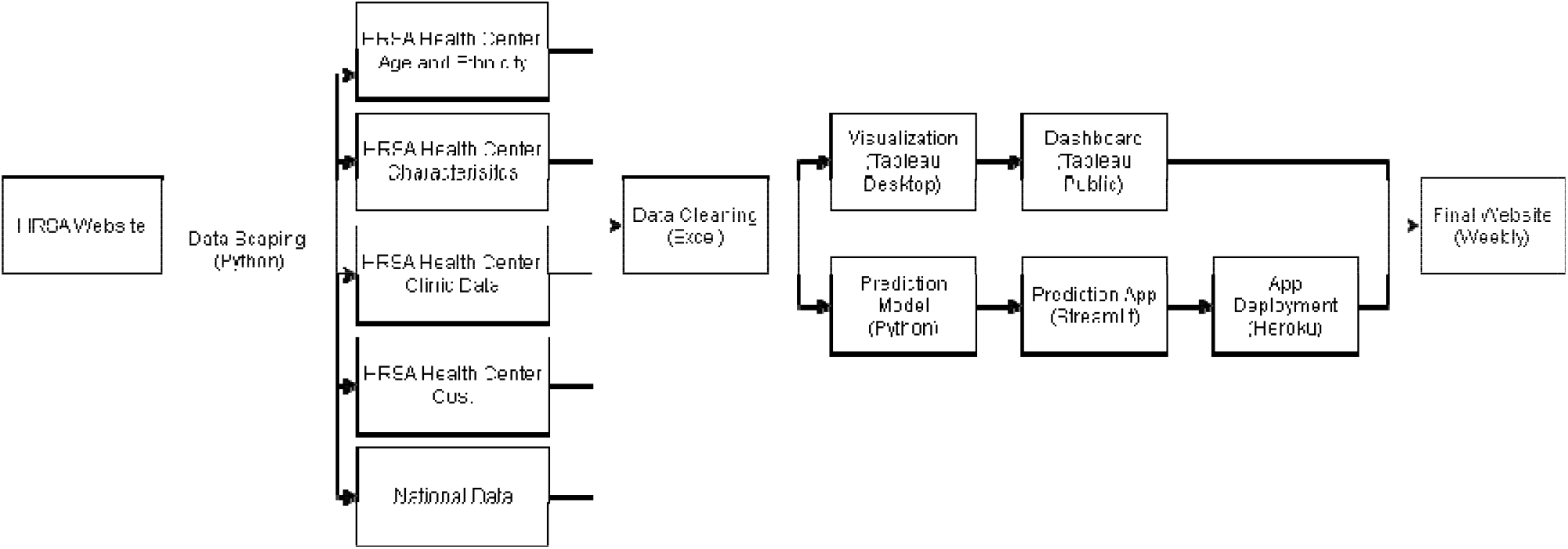
Dashboard Application System Design

## Results

There are two deliverables in this project. The data visualization is consolidated into 3 dashboard views: Effectiveness of Preventive Intervention, Effectiveness of Chronic Disease Management, and Financial Analysis. Each of the views allows users to choose the specific chronic disease management and preventive intervention program at each year and compare its performance with the disease prevalence and cost benefits. The dashboard focuses on showing the changes over the years to observe any improvement. Figure 3 shows the effectiveness of preventive intervention dashboard which compares the screening program rates with the average cost per patient at each state. The dashboard shows the measures in both map view and stacked bar view.

**Figure 3.**
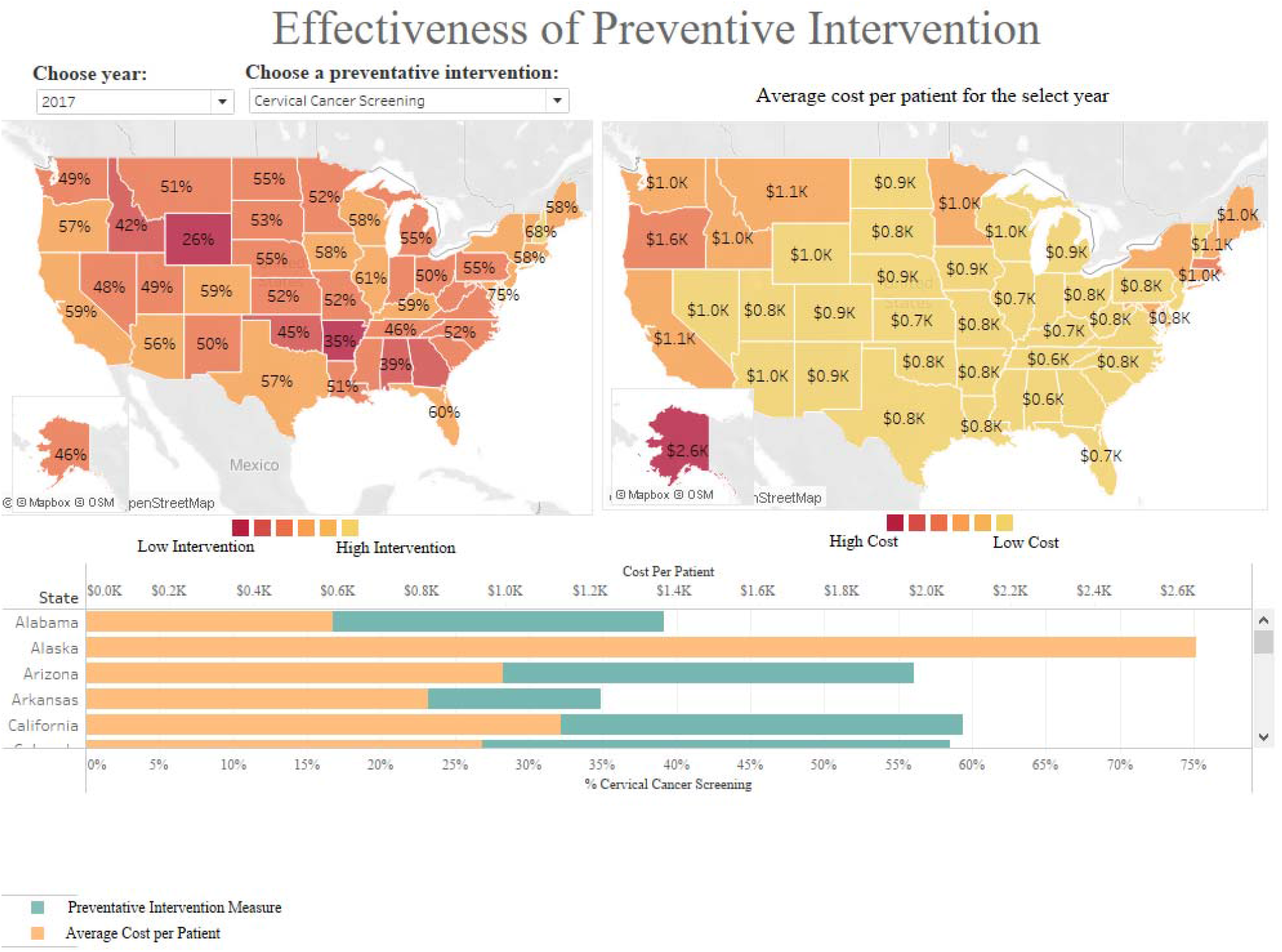
Effectiveness of Preventive Intervention

Figure 4 shows the effectiveness of chronic disease management by comparing the disease prevalence with the disease management programs. It also includes the national rate of disease prevalence and lists the cities that underperformed the national benchmark.

**Figure 4.**
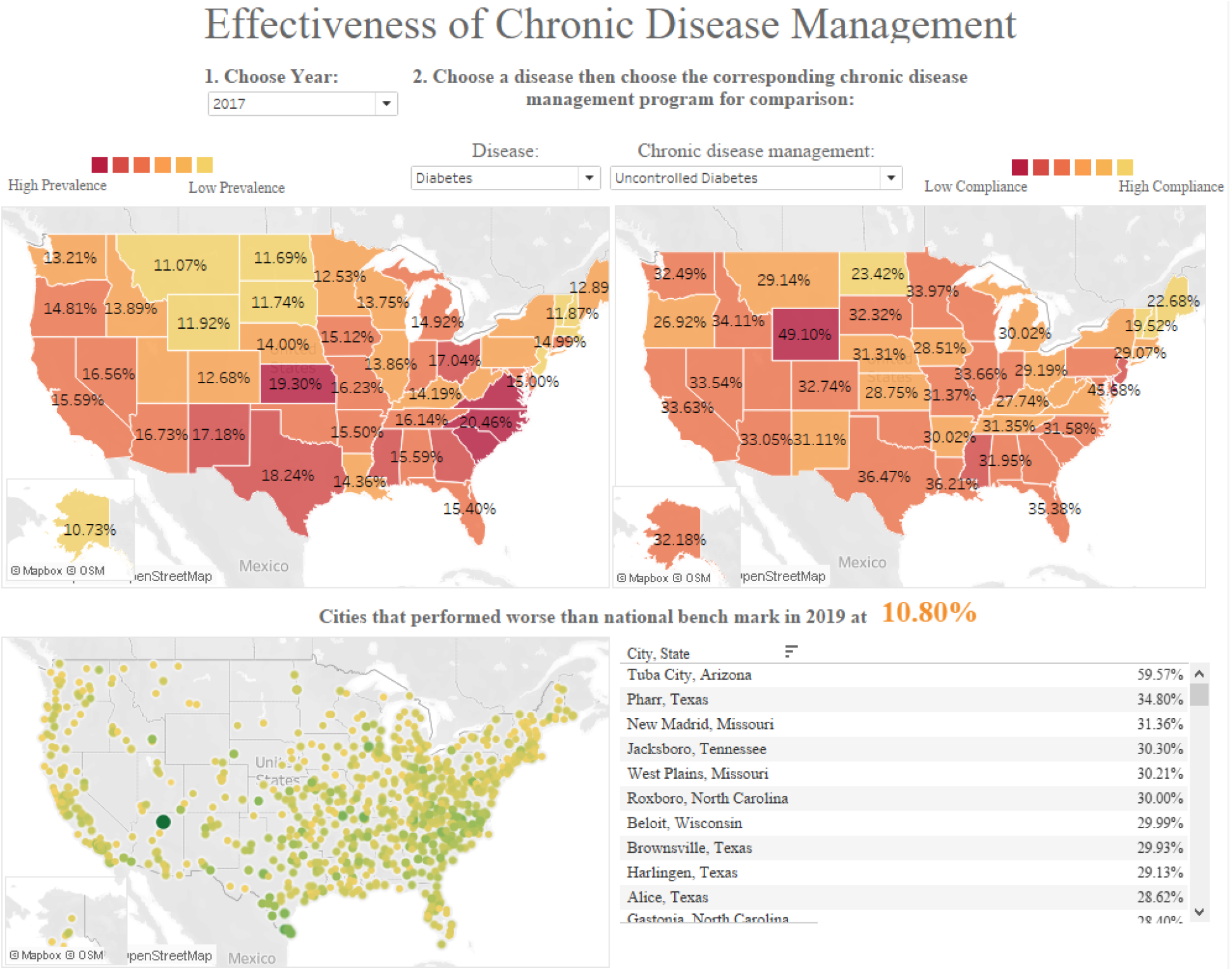
Effectiveness of Chronic Disease Management

Figure 5 shows the HRSA Financial Dashboard which highlights the cost per patient at each state compared to the national rate and cost categories. The dashboard also exhibits the charges and collections by different types of payers.

**Figure 5.**
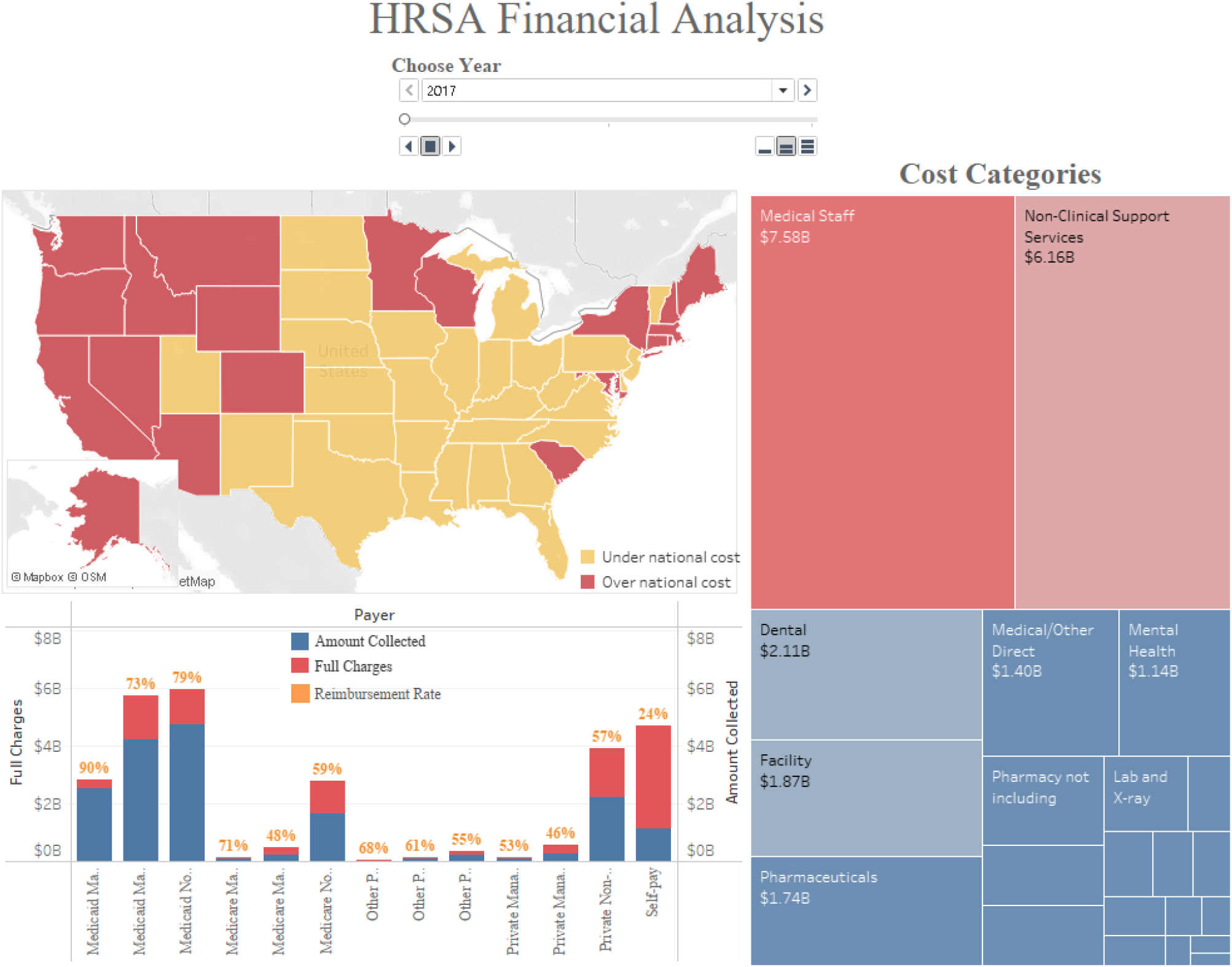
HRSA Financial Analysis

To design a prediction model based on HRSA-funded health center data, the project tested 10 different regression models available in scikit-learn Python library. Mean Squared Error (MSE) was chosen as the performance metric for the regression models. Based on the comparison in Figure 6, Extra Trees model outperformed all the other regression models with the smallest value of MSE. The Root Mean Squared Error (RMSE) of Extra Trees model was 409.05.

**Figure 6.**
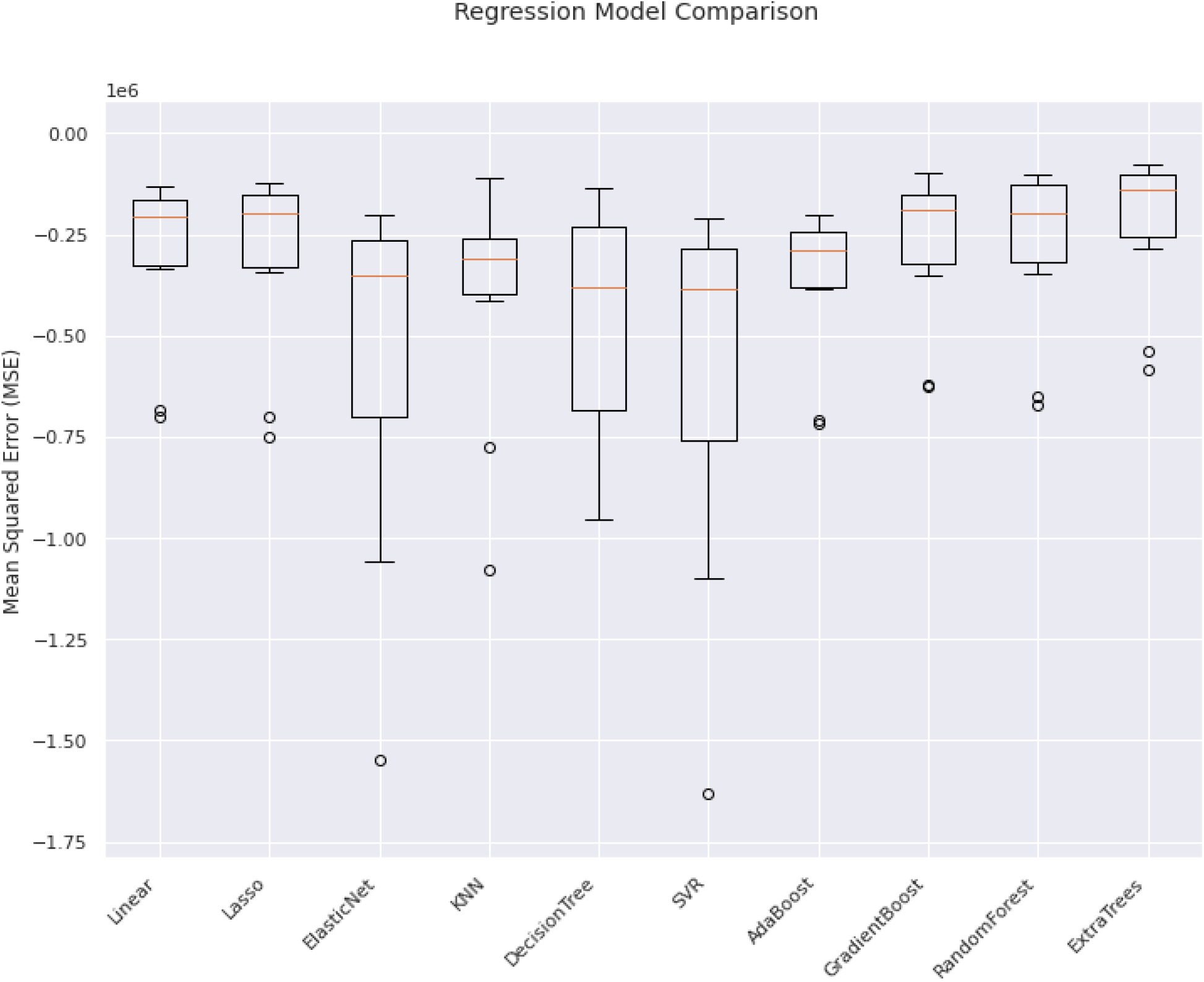
Regression Model Comparison

In addition, the project also examined the most important features that impact the prediction of healthcare cost. The feature importance function in Python library indicated that the rate of HIV patients, rate of patients receiving mental services, rate of minority patients, rate of veterans, rate of homeless patients, and rate of patients under 200% poverty line made the most impact on the cost prediction (see Figure 7). The model selection and feature selection were used to train the model. Finally, the Python model was deployed to become a user-interactive application.

**Figure 7.**
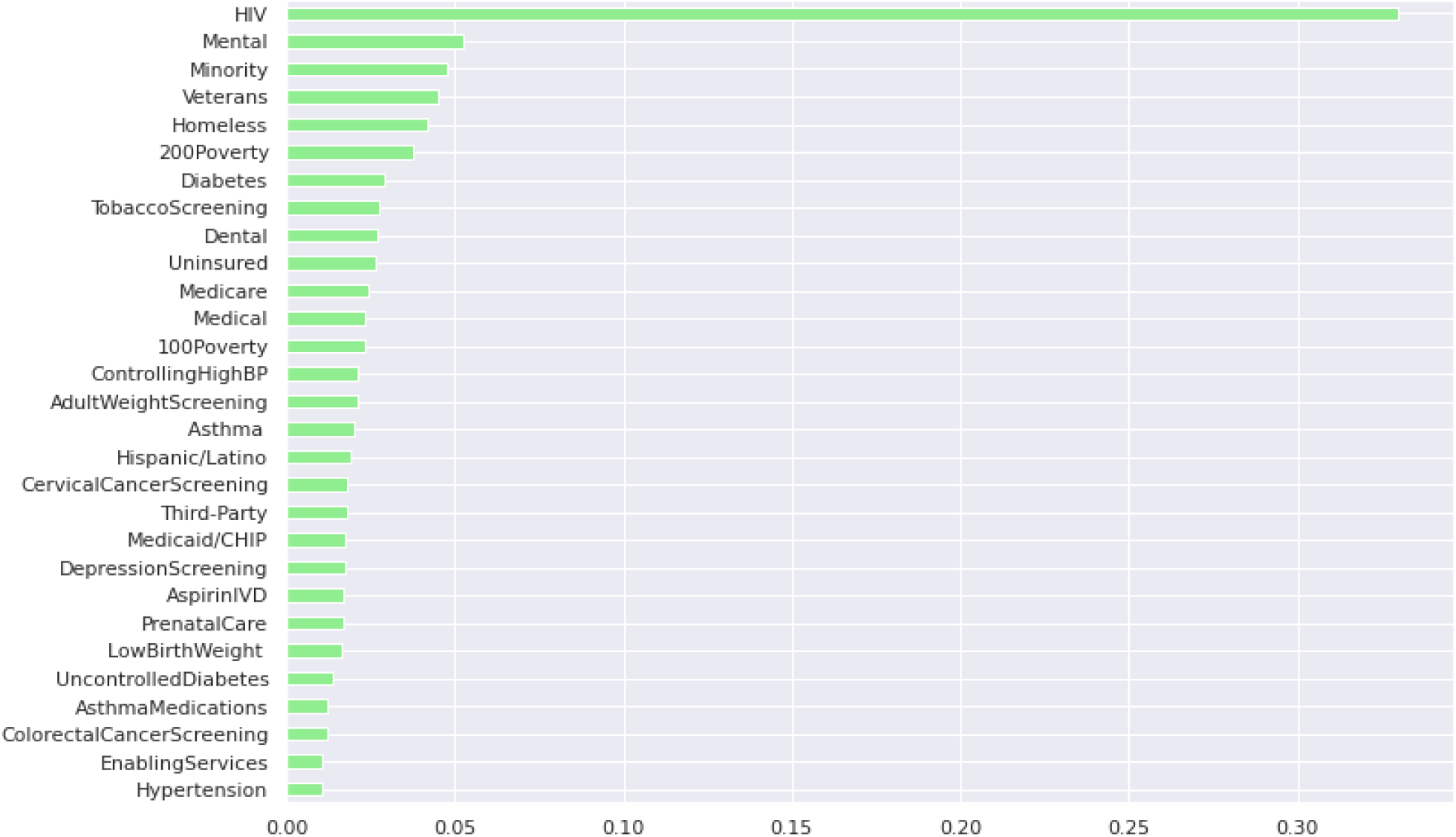
Feature Importance Selection

As shown in Figure 8, the final product allows selection of a specific value for each measure among the most important features and calculates the predicted cost per patient for that health center.

**Figure 8.**
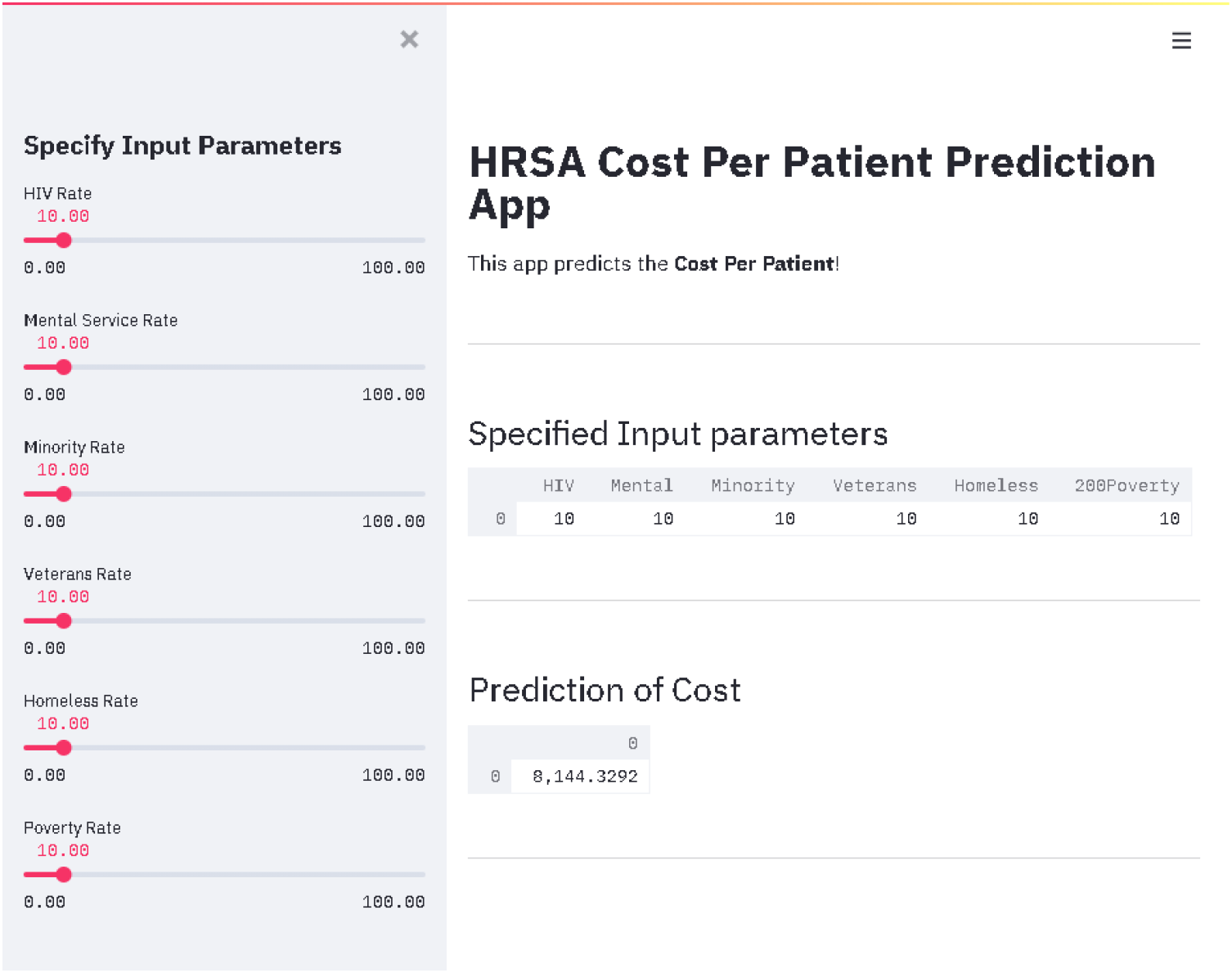
Cost Prediction Application

## Discussion

This design of the HRSA Population Health Dashboard will resolve the lack of performance comparison and trendline visualization among available HRSA dashboards. Such dashboard layout could generate meaningful insights for state- and city-level strategy-making.

The HRSA data from 2017 to 2019 is indicating the ineffectiveness of the preventive and chronic care programs in reducing disease prevalence and healthcare cost among states. Decision makers will need to investigate further why the more the prevention and management program they run, the higher the cost and disease prevalence rate are. The question raised is whether these screening and management programs benefit the long-term savings for the patients and health centers.

However, some limitations on running this HRSA data-driven website should be noted. HRSA only releases 3 years of data at a time so the current version of the dashboard is limited. HRSA data is in a difficult format to retrieve, thus extra time and effort are required to capture the full data set. There are also changes in clinical measure definitions over the years, which might impact the accuracy of visualization and prediction. Lastly, there are newly introduced measures that do not have enough data to trend and analyze.

### Next Steps/ Opportunities

The HRSA Population Health Dashboard is available for public use at the website https://hrsadashboard.weebly.com. The data behind this dashboard can be expanded further to include other information such as healthcare utilization, workforce, and payment to incorporate more factors into the comparison and trend analysis. We can continue to collect data every year to increase the accuracy of this prediction model. The website can have its user interface and user experience enhanced. Finally, there are HRSA-provided Programming Application Interfaces (APIs) that can connect the dashboard to real-time data update.

## Data Availability

All data produced are available online at

https://bphc.hrsa.gov/datareporting/reporting/index.html

## Notes

### Competing Interest Statement

The authors have declared no competing interest.

### Funding Statement

This study did not receive any funding

### Author Declarations

The study used (or will use) ONLY openly available human data that were originally located at: https://bphc.hrsa.gov/datareporting/reporting/index.html

